# Dietary Trends and Dementia - A Multi-Country Ecological Analysis

**DOI:** 10.1101/2023.01.09.23284344

**Authors:** Robert Reed

## Abstract

**Background:** This research evaluates the association between increased animal product consumption and population-level dementia incidence.

**Design/methodology/approach:** Publicly available data from 54 countries across a 10-year span was used to conduct a multivariate panel data regression to determine significant relationships between dietary changes and rates of dementia. Fixed effects analysis controls for the effect of omitted time-invariant differences between countries.

**Findings:** Excess calorie consumption is associated with a significant increase in dementia whereas fish consumption appears protective. On a population-level basis, increases in milk and egg consumption were associated with an increase in dementia rates. Meat, as a broadly defined category, was found to have no significant effect.

**Practical Implications:** This study supports the well-documented benefits of calorie control and fish consumption to protect against dementia, but indicates that egg and milk consumption is associated with increased population-level dementia incidence.

**Originality/value:** This research expands current literature by using an updated data set, evaluating considerably more countries, and utilizing a regression model that controls for the effects of time-invariant sources of heterogeneity in the data.

## Introduction

Dementia is typically defined as a persistent clinical syndrome of cognitive decline that interferes with the ability to remember, think, make decisions, and has a substantive impairment on the ability to perform everyday tasks (Chertkow *et al*., 2013). In addition to dementia’s personal toll on patients and their loved ones, there is a substantial economic burden with annual expenditures of €32,000 per patient in Europe and over €42,000, in the United States (Cantarero-Prieto *et al*., 2020). In 2018, the total estimated burden of Alzheimer’s disease and related dementias was $2.8 Trillion with a projected increase to $16.9 Trillion by 2050 (Nandi*et al*., 2022).

While Langa (2015) does find some evidence of decreasing dementia rates in the West, the actual number of affected individuals is projected to increase with the most rapid increases of 367% and 357% occurring in North Africa and the Middle East and eastern sub-Saharan Africa respectively (Nichols *et al*., 2022). Despite the momentous effort to find a treatment for this costly and devastating disease, Poudel and Park (2022) highlight that existing treatments show limited efficacy and are limited to providing symptomatic solutions. This makes preventing dementia a pressing medical, social, and economic challenge.

Given the immense cost of dementia and the invariably fatal outcome of a dementia diagnosis, Fessel (2021) asserts that prevention is better than a cure, and books such as Dan Buettner’s 2008 *The Blue Zones*, which suggest that low-cost dietary modifications, such as limiting the consumption of animal products, can delay, slow, or reduce the population level incidence of dementia, have understandably received both enthusiasm and considerable attention from physicians, economists, and public health researchers alike.

Zissimopoulos *et al*. (2014) estimate that even delaying the onset of Alzheimer’s by 5 years could lead to a 41% lower prevalence and 40% lower cost by 2050. If the proposed benefits of such plant-based diets were shown to have even a moderate protective effect, the economic and societal benefits would be impressive.

Despite the appeal of a dietary strategy for reducing dementia incidence, research has yielded mixed results. Although prevailing dietary wisdom (Quan *et al*., 2022) indicates that lowering meat consumption may be beneficial for dementia outcomes, this is not universally accepted with Ylilauri *et al*., (2022) finding that there is no association between animal product consumption and dementia. Other studies have found protective effects of egg (Lee *et al*., 2021), whole milk (Muñoz-Garach *et al*, 2021), and meat consumption (Ngabirano *et al*., 2019).

Even the remarkable health outcomes of the so-called Blue Zones themselves have been called into question, thereby obfuscating the purported benefits of the underlying diet. Newman (2019) postulates that the health outcomes of the Blue Zones come not from the diet, but rather from record-keeping errors, and in some cases, the link between animal consumption and Alzheimer’s vanishes when accounting for changes in diagnostic criteria (Wu *et al*., 2015). Rosero-Bixby (2013) found that the theorized benefit of the Nicoya Blue Zone applied only to men. Gavrilova and Gavrilov (2012) contend that whatever dietary benefit may have once existed is quickly fading as the longevity benefit in the Okinawan Blue Zone only applies to those born prior to World War II; the life expectancy at birth for men in Okinawa is now lower than the country average.

To derive a more conclusive relationship between diet and population-level dementia incidence, we must expand the scope of research beyond individual trials or even communities and analyze trends at the country level. Grant’s (2014) study demonstrated that Japan’s increased animal fat and calorie consumption was highly correlated with increased AD prevalence, but confirmation of this finding, and the expansion of the study to include more countries, is needed to yield further insight.

This study seeks to expand the current body of population-based research by using a panel data analysis comprising 54 countries across a 10-year span to evaluate the link between dietary trends and dementia incidence.

## Methods

Data collection was performed in a two-stage collection and cleaning process. In the initial collection phase, the age-standardized death rate per 100,000 from dementia was obtained from the WHO’s “Alzheimer and other dementias” mortality database, while meat, milk, egg, pelagic fish, and total calorie consumption was gathered from the Food and Agriculture Organization of the United Nation’s FAOSTAT database.

Once initial data was collected, Python’s Pandas library was used to successively merge the datasets into a complete dataset by matching data by country and year. Automating the data merging in this way reduces the possibility of human error in manually aligning columns, however, it does mean that countries that changed names during the course of the data period or are reported differently will be dropped from the data. Lastly, any country/year pairs that included null values were dropped from the dataset for that specific year.

To protect the model from the impact of improper disease estimates, the data was further refined to include only the countries that met the WHO’s “high quality” standard for data reporting. FAOSTAT did not differentiate food supply estimates based on the quality of data. In the absence of an FAOSTAT disclaimer that certain countries did not meet the quality of data reporting, it was presumed that data were valid. Further, it is likely that countries that met the WHOs high-quality standards for disease reporting would likewise have adequate infrastructure and policies to facilitate an equally high standard of food reporting.

After all data cleaning was performed, there were a total of 54 countries represented in the study with data spanning 2010 to 2019 for a total of 479 unique observations. Due to data unavailability for some years, not all countries were represented in all time periods.

A fixed effects panel data regression model (FE model) was identified as the most appropriate model for the data because it allows the researcher to estimate the impact of each independent variable (IV) on the dependent variable (DV) of Alzheimer’s rate while holding the effects of other variables constant. Because the FE model uses both cross-sectional data (multiple countries) and time series data (each country across multiple years), it is able to treat each country as its own control thereby minimizing the effects of time-invariant sources of heterogeneity between countries. In other words, the model can still report accurate IV/DV relationships when presented with countries that have different cultural attitudes toward aging or different leisure preferences so long as these preferences remain relatively stable over time. This assumption is discussed further in the limitations.

### Statistical Analysis

The FE model was conducted with gretl version 2022a with unit data indexed by country name and time data indexed by year. Age-standardized Alzheimer’s rate was used as the DV with calories, milk, meat, eggs, and pelagic fish consumption as IVs. As shown in Table I, increased milk, egg, and total calorie consumption showed a positive association with age-standardized dementia incidence. Pelagic fish consumption (which includes tuna, mackerel, herring, etc) showed an inverse relationship with dementia rates. An inverse relationship between meat and dementia rates fell shy of meeting the .05 level of significance but still bears further discussion.

**Table I:** Regression output and p-values.

| Associations Between Diet and Age-Adjusted Alzheimer's Rate |  |  |  |  |
| --- | --- | --- | --- | --- |
|  | coefficient | std. error | t-ratio | p-value |
| const | -22.467 | 7.528 | -2.985 | 0.004*** |
| Calories | 0.010 | 0.003 | 3.594 | 0.001*** |
| Milk | 0.028 | 0.009 | 3.048 | 0.004*** |
| Meat | -0.056 | 0.031 | -1.806 | 0.077* |
| Eggs | 0.557 | 0.172 | 3.244 | 0.002*** |
| Pelagic | -0.346 | 0.133 | -2.595 | 0.012** |

### Discussion

Notwithstanding the concern of mercury (Storelli, 2008), fish are generally accepted as neuroprotective and are included as a key component of both the Mediterranean (Guasch-Ferré & Willett, 2021) as well as MIND diet (Morris *et al*., 2015). In accordance with prevailing studies that find a protective effect of fish consumption (Roberts *et al*., 2012), the data in this research revealed an inverse relationship between population-level fish consumption and dementia incidence. On the balance, it appears the benefits of fish consumption outweigh the risks (Mozaffarian & Rimm, 2006).

This study’s finding of a positive link between population-level consumption of dairy and dementia supports the findings of Muñoz-Garach*et al*., (2021) who noted higher Mini-Mental State Evaluation (MMSE) scores among those who consumed less milk. It also provides support for Petruski-Ivleva *et al*., (2017) who identified a positive association between midlife milk consumption and dementia. Although a recent meta-analysis by Lee *et al*., (2018) concluded that the balance of evidence is not sufficient to suggest a causal relationship between dairy consumption and dementia, the positive link found in this study, combined with the aforementioned research, suggests a more nuanced study of the nature between dairy and dementia may prove worthwhile for elucidating the full role that milk may play.

Although Tsai (2015) found negative cognitive outcomes when eggs were consumed as part of a “Western Diet”, egg-specific studies have reported a neutral (Bishop & Zuniga, 2019) or in other cases protective (Lee *et al*., 2021) relationship with dementia. The neuroprotective effect of choline from eggs is widely documented (Ylilauri et al., 2019; Yuan et al., 2022). However, the detrimental effect of egg consumption found in this study should not be dismissed out of hand as there is a theoretical connection between saturated fat (Ruan *et al*., 2018) and dietary cholesterol (Kalmijn *et al*., 1997) found in eggs and increased dementia risk.

Based on 2016 estimates by Tc and Vl, 90% of the US population is choline deficient which could mean that at low to moderate doses, the protective effect of choline outweighs the negative saturated fat and cholesterol of eggs. After a certain egg intake helps individuals achieve choline sufficiency, it is possible that additional intake past this point carries only increased risk via cholesterol and sat fat. Although the high-level nature of this data set precludes further investigation, more nuanced studies could reveal additional information regarding the interaction between egg consumption and dementia incidence when adjusting for moderating variables including external choline sources.

The non-significant impact of meat in this study aligns with the unsettled nature of the question in the extant literature. Ngabirano *et al*., (2019) found that low meat consumption was associated with increased dementia risk. Likewise, processed meat has been found to be detrimental, but unprocessed red meat has been shown to be protective (Zhang *et al*., 2021). The data in this study did not differentiate between types of meat or preparation methods, so it is possible that the protective effects of some meats and the detrimental effects of others counterbalanced each other to produce an insignificant finding. The insignificant findings of this study are unable to provide support for or against meat, but rather echo calls for further research to disentangle the complex relationship that different types of protein sources and preparation methods may have with dementia incidence (Ylilauri *et al*., 2022)

### Limitations

Although necessary to ensure the accuracy of the research, the data filtering and preparation process reduced the breadth of data available for study. While the sample size was sufficiently large and diversified to draw meaningful conclusions, the inclusion of more countries with diverse dietary habits could have yielded additional insight had the data been available.

Diet is just one of countless factors that could influence the reported Alzheimer’s rate in a country. Whereas a traditional regression model only accounts for the independent variables that are explicitly included in the model, the FE model used in this analysis does control for omitted variables between countries so long as those omitted factors are stable through the course of the study. For example, the model can account for the effect of different cultural attitudes toward aging or access to healthcare between countries so long as these differences are chronic.

The ability to account for the time-invariant sources of heterogeneity among countries is a strength of the FE model, but it is important to remember that even the most well-designed FE model is still a simplification of reality that can not capture real–time sources of variation. Though powerful, significant variation will remain unexplained by the model, and the findings here should not be used as “proof” of causation. Rather the findings should be analyzed within the context of providing additional clarity and context to existing research and theory.

## Conclusion

Dementia is a progressive, incurable disease that has massive health, economic, and social costs that are only projected to increase in the coming years. The limited effectiveness of standard medical treatment makes the prevention of dementia through dietary factors an appealing proposition with numerous diets claiming to offer neuroprotective effects by limiting the intake of animal products. Where small-scale studies have yielded conflicting results, this study seeks to analyze large-scale trends between diet and dementia at the population level.

As with all observational studies, this research is incapable of proving a causal relationship, and poor quality data from some countries meant that the researcher was forced to limit the breadth of countries included in the sample in favor of preserving the validity of the results. Despite the limitations of the study, this multi-country longitudinal analysis finds meaningful, population-level associations between dietary trends and age-standardized dementia incidence. The detrimental effect of excess calories and the protective effect of fish consumption align with both existing research consensus as well as recommendations of the MIND, Mediterranean, and Blue Zone Diets. Though existing research is less decisive on the matter, this study does find a positive association between dementia incidence and the consumption of eggs and milk.

In practical terms, this research suggests the population-level benefits of various diets and public health initiatives that promote fish consumption and moderated caloric intake. Where existing research has yet to reach a consensus, this study indicates the possibility of a positive association between dementia incidence and the consumption of milk and eggs.

## Data Availability

All data is publicly available from the WHO and UN

https://www.fao.org/faostat/en/#data/FBS

https://platform.who.int/mortality/themes/theme-details/topics/indicator-groups/indicator-group-details/MDB/alzheimer-and-other-dementias

